# Schizophrenia-associated differential DNA methylation in the superior temporal gyrus is distributed to many sites across the genome and annotated by the risk gene *MAD1L1*

**DOI:** 10.1101/2020.08.02.20166777

**Authors:** Brandon C. McKinney, Christopher M. Hensler, Yue Wei, David A. Lewis, Jiebiao Wang, Ying Ding, Robert A. Sweet

## Abstract

**Background:** Many genetic variants and multiple environmental factors increase risk for schizophrenia (SZ). SZ-associated genetic variants and environmental risk factors have been associated with altered DNA methylation (DNAm), the addition of a methyl group to a cytosine in DNA. DNAm changes, acting through effects on gene expression, represent one potential mechanism by which genetic and environmental factors confer risk for SZ and alter neurobiology.

**Methods:** We investigated the hypothesis that DNAm in superior temporal gyrus (STG) is altered in SZ. We measured genome-wide DNAm in postmortem STG from 44 SZ subjects and 44 non-psychiatric comparison (NPC) subjects using Illumina Infinium MethylationEPIC BeadChip microarrays. We applied tensor composition analysis to extract cell type-specific DNAm signals.

**Results:** We found that DNAm levels differed between SZ and NPC subjects at 242 sites, and 44 regions comprised of two or more sites, with a false discovery rate cutoff of q=0.1. We determined differential methylation at nine of the individual sites were driven by neuron-specific DNAm alterations. Glia-specific DNAm alterations drove the differences at two sites. Notably, we identied SZ-associated differential methylation within within mitotic arrest deficient 1-like 1 (*MAD1L1*), a gene strongly associated with SZ through genome-wide association studies.

**Conclusions:** This study adds to a growing number of studies that implicate DNAm, and epigenetic pathways more generally, in SZ. Our findings suggest differential methylation may contribute to STG dysfunction in SZ. Future studies to identify the mechanisms by which altered DNAm, especially within MAD1L1, contributes to SZ neurobiology are warranted.

## INTRODUCTION

Schizophrenia (SZ) is a severe neuropsychiatric disorder with complex etiology. Heritability estimates for SZ from twin studies are consistently ∼80% (1), thus suggesting a substantial genetic contribution to its etiology. Genome-wide association studies (GWAS) have identified many common variants associated with SZ, although each variant has only a small effect on risk for the disorder (2). A recent large-scale GWAS meta-analysis identified such variants at 145 distinct genetic loci (3). Heritability estimates from GWAS fall short of those predicted by twin studies, thus suggesting that other forms of genetic variation contribute risk for SZ. Indeed, recent studies have found a high burden of both rare single nucleotide variants and rare copy number variants in subjects with SZ (4,5). Multiple environmental factors have also been found to contribute to SZ etiology including, among others, maternal infections and malnutrition during pregnancy and obstetric complications (6).

SZ-associated genetic variation and environmental factors associated with SZ risk have both been found to alter DNA methylation (DNAm) (7–11). DNAm, the addition of a methyl group to a cytosine in DNA, stably affects gene experession via interaction with transcription factor binding (12). DNAm is associated with both increased and decreased gene expression as well as other forms of gene regulation, including splicing and alternative promoter usage (12–14). Changes in DNAm, acting through effects on gene expression, represent one potential mechanism by which both genetic and environmental factors can confer risk for SZ and alter brain development and biology.

The superior temporal gyrus (STG) is a region of the brain critical for auditory processing. In individuals with SZ, altered STG function is associated with auditory verbal hallucinations and impaired auditory sensory processing. Impaired auditory processing further contributes to phonologic dyslexia and difficulty recognizing and expressing spoken emotional tone (prosody) in SZ (15). In this study, we investigated the hypothesis that DNAm is altered in the STG from subjects with SZ. To this end, we used Illumina Infinium MethylationEPIC BeadChip microarrays (EPIC arrays) to measure genome-wide DNAm in the STG from 44 subjects with SZ and 44 non-psychiatric comparison (NPC) subjects. Tensor composition analysis (TCA) was used to extract cell type-specific DNAm signals from brain tissue-level data.

## MATERIALS AND METHODS

### POSTMORTEM BRAINS

Tissue was obtained from postmortem brains recovered and processed as described previously (16,17). Briefly, brains were retrieved during routine autopsies at the Allegheny County Medical Examiner’s Office, Pittsburgh, PA, USA, following informed consent from next-of-kin. An independent committee of experienced clinicians made consensus Diagnostic and Statistical Manual of Mental Disorders, Fourth Edition diagnoses, or determined the absence thereof, based on clinical records and collateral history obtained via structured interviews with surviving relatives (18). The right hemisphere was blocked coronally and the resultant slabs snap frozen and stored at −80°C. Slabs containing the STG were identified and the STG was removed as a single block from each of the slabs in which it was present. Samples containing all six cortical layers of STG (planum temporale), but excluding the adjacent white matter, were harvested. All procedures were approved by the University of Pittsburgh Committee for the Oversight of Research and Clinical Training Involving Decedents and the Institutional Review Board for Biomedical Research.

### COHORT MEMBERSHIP

The cohort comprised 44 subjects with either schizophrenia (N=31) or schizoaffective disorder (N=13), and 44 NPC subjects. Subjects diagnosed with schizophrenia and schizoaffective disorder were grouped together for analysis, and referred to as SZ subjects, or the SZ group. In this manuscript, as in our previous studies, we found that the diagnoses do not differ with respect to DNAm (19). Each subject in the SZ group was matched with one NPC subject for sex, hemisphere, and as closely as possible for postmortem interval (PMI), age, and other characteristics (Table 1 and Supplementary Table 1).

**Table 1.**
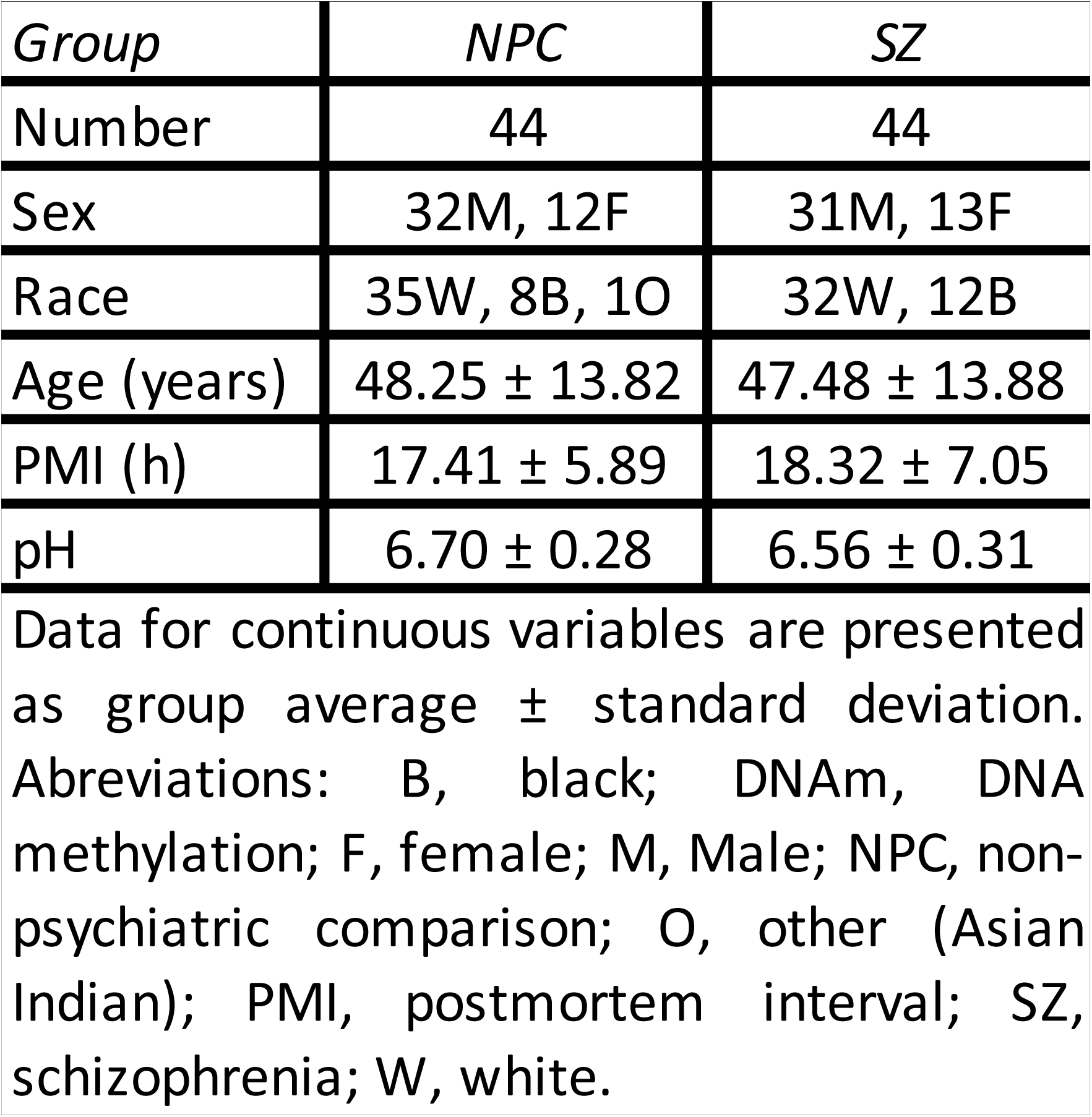
Cohort characteristics.

### DNA PREPARATION AND BISULFITE CONVERSION

DNA (∼10 ug) was isolated from STG gray matter (∼20 mg) using AllPrep DNA/RNA/Protein Mini Kit (Qiagen, Valencia, CA, USA) and bisulfite converted using EZ-96 DNA Methylation Kit (Zymo Research, Irvine, CA, USA), both as per manufacturer’s protocol.

### DNA METHYLATION ARRAYS

DNAm is the addition of a methyl group to a cytosine in DNA. DNAm is observed within the context of cytosine-phosphate-guanine dinucleotides (CpGs), most commonly, but also within the context of a cytosine-phosphate-H dinucleotides (CpHs; H=cytosine, adenine, thymine) (20,21). CpGs and CpHs are referred to as ‘DNAm sites’ or ‘sites’ in this manuscript. DNAm was measured at 866,091 sites using MethylationEPIC BeadChip Infinium array (EPIC array; Illumina, San Diego, CA, USA) as per manufacturer’s protocol (22,23). A *β*-value, the proportion of a particular site that is methylated in a DNA sample, was determined for each site by taking the ratio of the methylated to unmethylated signal, using the formula: *β*-value = intensity of the methylated signal/(intensity of the unmethylated signal + intensity of the methylated signal + 100). A 96-entry EPIC array was filled with samples from the 88 subjects, including replicate samples from eight subjects. Data is available for download from Gene Expression Omnibus (GEO; GSE144910).

### DATA PREPROCESSING AND FILTERING

Data analyses were performed using the R software environment (www.r-project.org). Color adjustment and background correction were performed using the bgAdjust2C method (24). Normalization was performed using the *preprocessQuantile* function in the R package *minfi* (25). The initial dataset comprised data from 1,051,815 probes corresponding to 866,091 DNAm sites for each subject. Multidimensional scaling (MDS) was used to visualize the degree of similarity among samples (26). Prior to data filtering, samples segregated strongly by sex in MDS space (Supplemental Figure 1A). Data filtering involved removing all data points associated with a probe if the probe failed detection as indicated by a median detection p-value >0.01 (probes corresponding to 12,350 DNAm sites), cross-reacted with multiple genomic regions (probes corresponding to 39,269 DNAm sites), contained a single nucleotide polymorphism within its binding site (probes corresponding to 27,395 DNAm sites), or interrogated a DNAm site on a sex chromosome (probes corresponding to 18,628 DNAm sites). Data from probes corresponding to 768,449 DNAm sites remained for downstream analysis (Supplemental Figure 2). After data filtering, MDS using data from the 3,000 most variable sites was performed and samples no longer segregated by sex (Supplemental Figure 1B), but segregation by race (Supplemental Figure 1C) and age (Supplemental Figures 1D and 1E) became evident. The replicate sample pairs from each of the eight subjects from which replicate samples were collected and assayed co-segregated in MDS space (Supplemental Figure 1F), thus demonstrating the reproduceability of our approach. The *β*-values for each replicate pair were averaged for the downstream analyses.

### DIFFERENTIAL DNA METHYLATION

Linear regression was used to identify differentially methylated sites (DMSs). DNAm, in the form of preprocessed and normalzed *β*-values, was the dependent variable and diagnosis was the independent variable. Race, age, and PMI were included as covariates in the analysis. The MDS analysis described above supported the inclusion of race as a covariate. Most subjects in this cohort self-identifed as either white or black, however, one subject self-identified as Asian Indian and, consistent with known genetic architecture (27), clustered with the subjects of European ancestry (Supplemental Figure 1C) and was thus combined with the subjects that self-identified as white for analyses. The inclusion of age as a covariate is supported by the MDS analysis as well as existing literature that shows age has a robust effect on DNAm (28–30). Though samples did not segregate by PMI in MDS space (data not shown), it was included as a covariate because the stability of many molecular measures have been found to be particularly sensitive to PMI (31,32), and to maintain consistency with our previous study in which it was included as a covariate in our primary analyses (19).

Differential methylated regions (DMRs) were identified using the R package DMRcate (33). DMRcate uses an approach based on tunable kernel smoothing of the differential methylation signal across the genome obtained in the site-based differential DNAm analysis described above. A Benjamini–Hochberg corrected false discovery rate (FDR) <0.1 for the smoothed signal was considered significant. Then regions with a maximum of 1000 basepairs containing at least two such significant sites were defined as DMRs.

### NEURON AND GLIA PROPORTION ESTIMATES

The proportion of neurons and glia in each sample was estimated with CETS, an R package that uses *β*-values from cell type-specific sites to generate the estimation (34).

### NEURON- AND GLIA-SPECIFIC DIFFERENTIAL DNA METHYLATION

The CETS-estimated proportions of neurons and glia for each subject and tensor composition analysis (TCA) (35) were used to estimate the subject-level neuron- and glia-specific *β*-values for each DNAm site and detect sites at which DNAm differs between subjects with SZ and NPC subjects. The cell type proportions were refit in TCA and adjusted for age, race, and PMI.

## RESULTS

### SZ-associated differential DNA methylation was identified at many individual sites and genomic regions

DNAm differed between subjects with SZ and NPC subjects at more sites than would be expected by chance (Figure 1A). DNAm differed at 242 sites between subjects with SZ and NPC subjects with an FDR cutoff of q=0.1 (Table 2). Of these 242 differntially methylated sites (DMSs), DNAm differed at 101 with a FDR cutoff of q=0.05. No global differences in DNAm were identified between SZ and NPC subjects (Supplemental Figure 3). The sites at which DNAm differed between subjects with SZ and NPC subjects were broadly distributed across all autosomes (Figure 1B).

**Figure 1.**
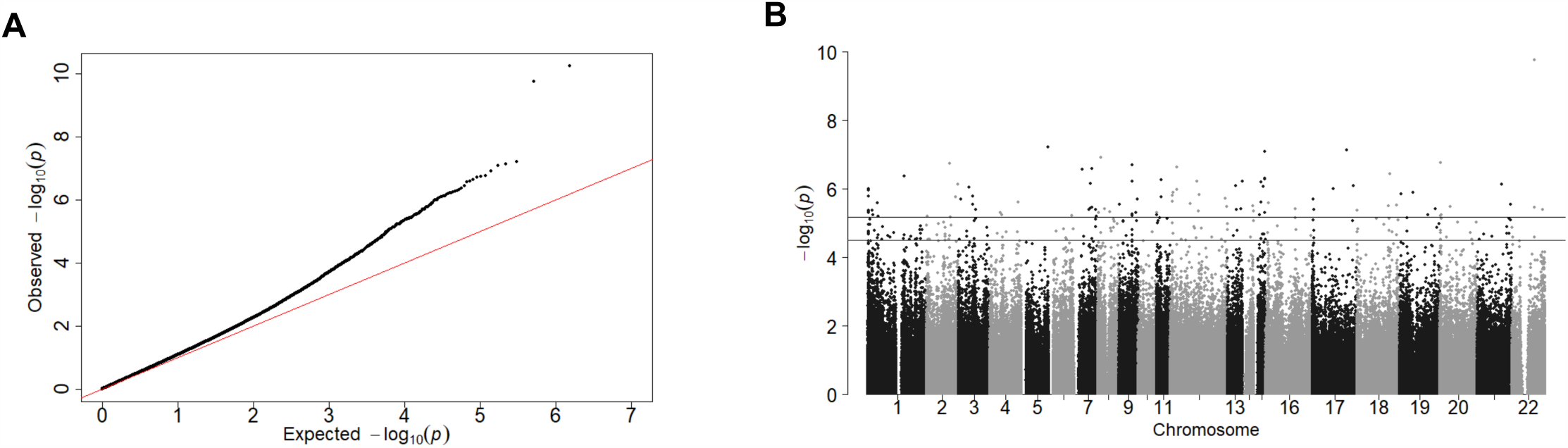
SZ-associated differential methylation. **(A)** Probability plot showing that the analysis for sites at which DNAm differed between SZ and NPC subjects is enriched in small p-values compared to what would be expected by chance. The y=x line represents the distribution of p-values than would be expected by chance. **(B)** Manhattan plot showing that the DNAm differed between subjects with SZ and NPC subjects at many DNAm sites, and the sites were distributed across many autosomes. The horizontal lines represent FDR cutoff of q=0.1 (bottom) and q=0.05 (top). DNAm, DNA methylation; SZ, schizophrenia; NPC, non-psychiatric comparison; FDR, false discovery rate.

**Table 2.**
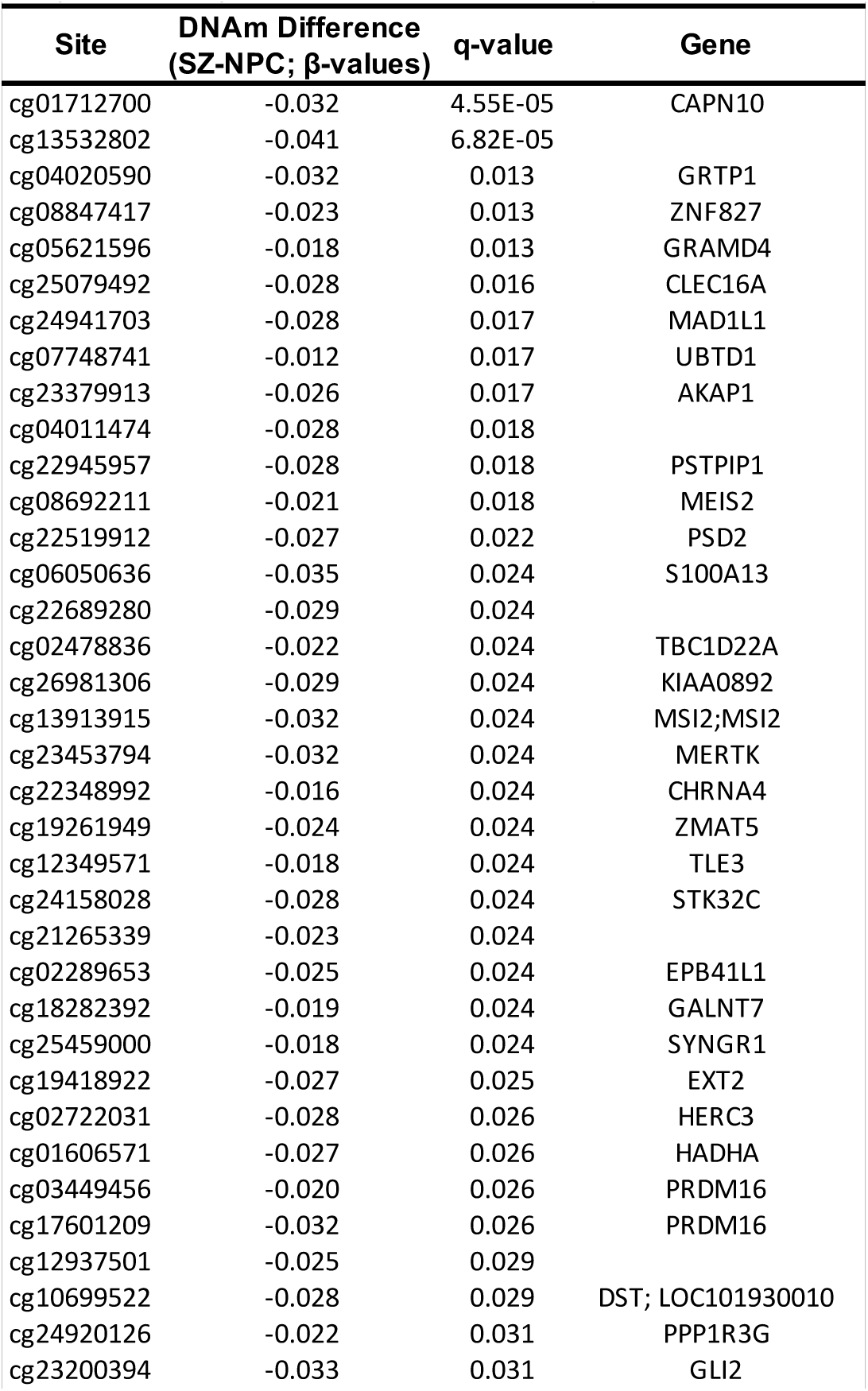
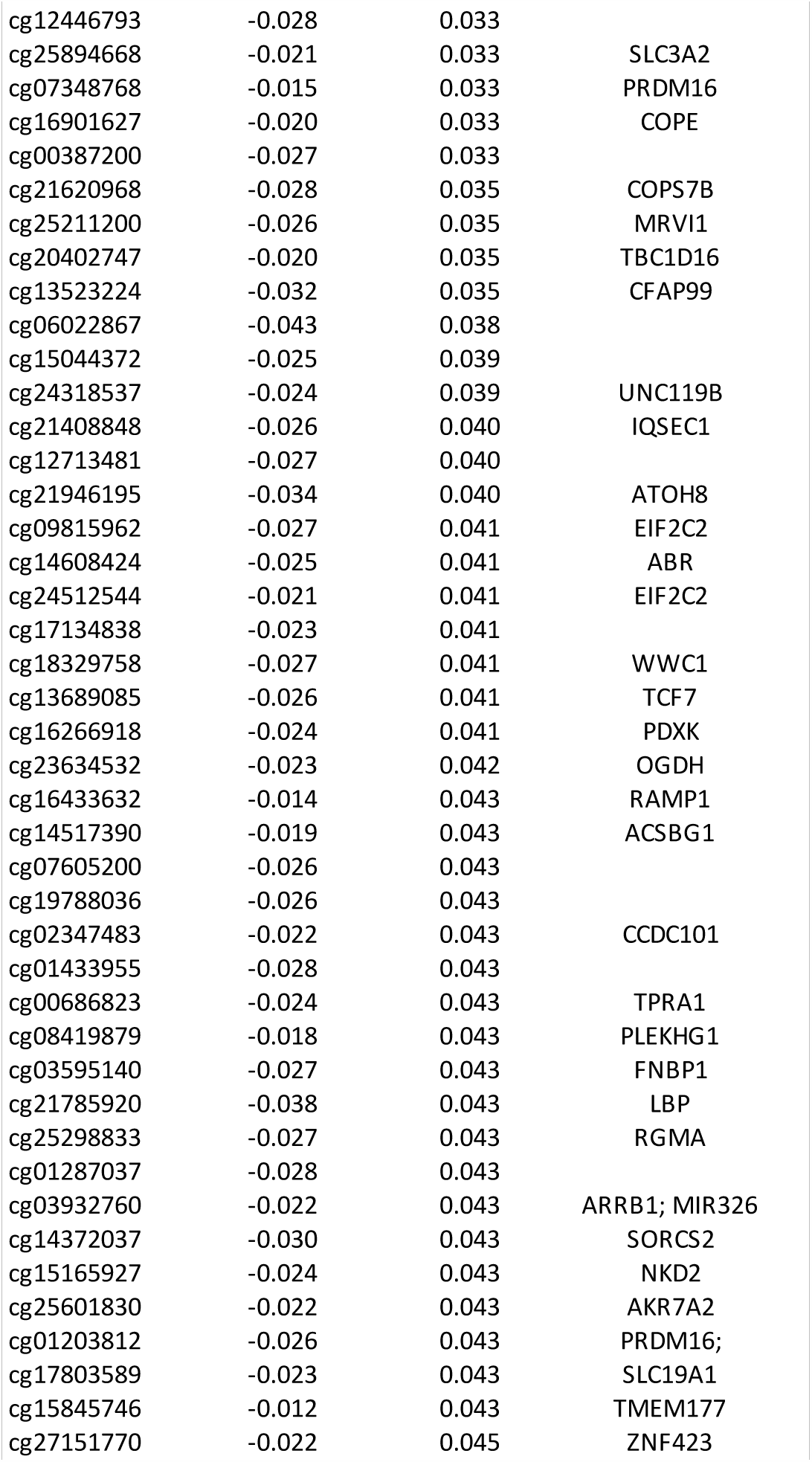
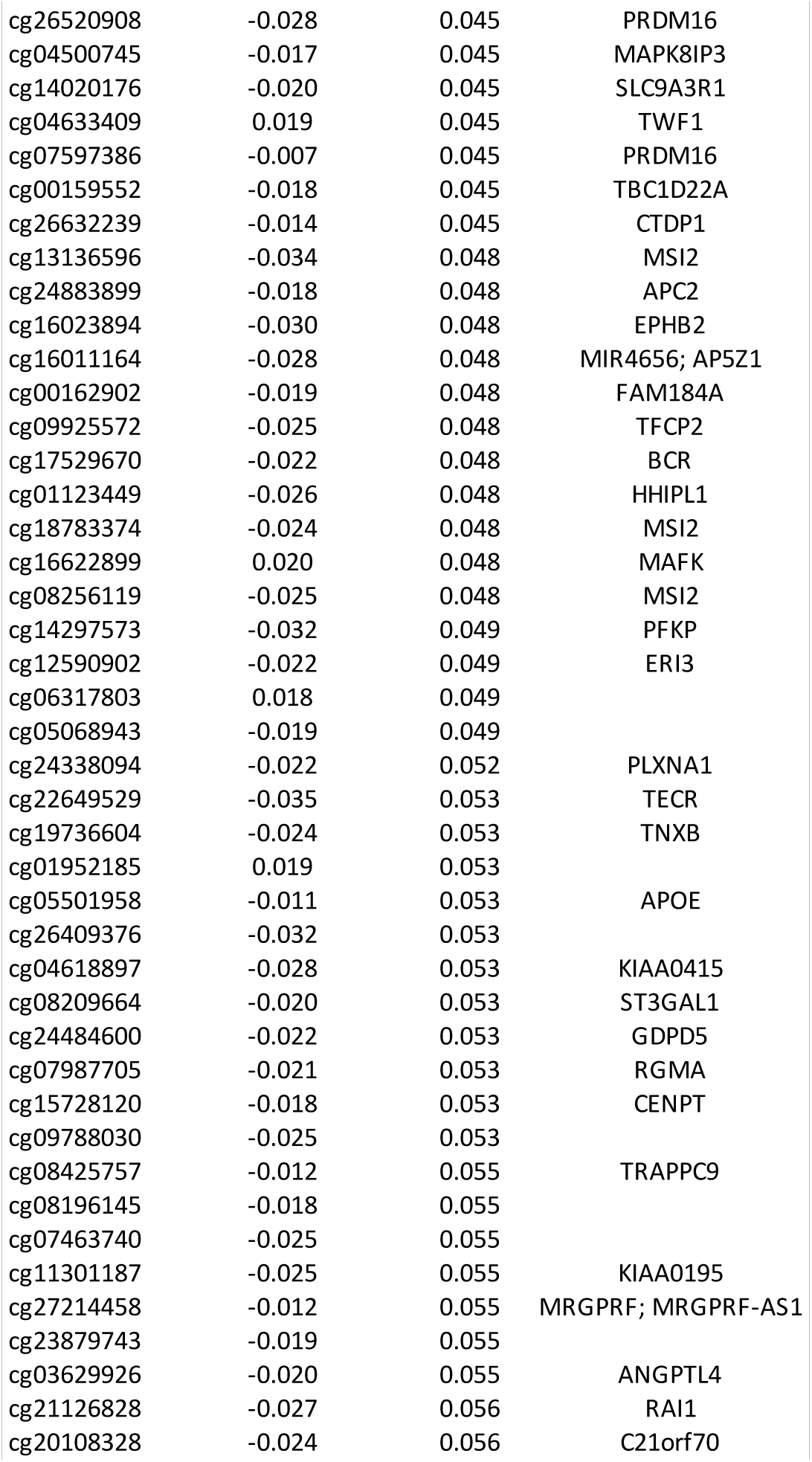
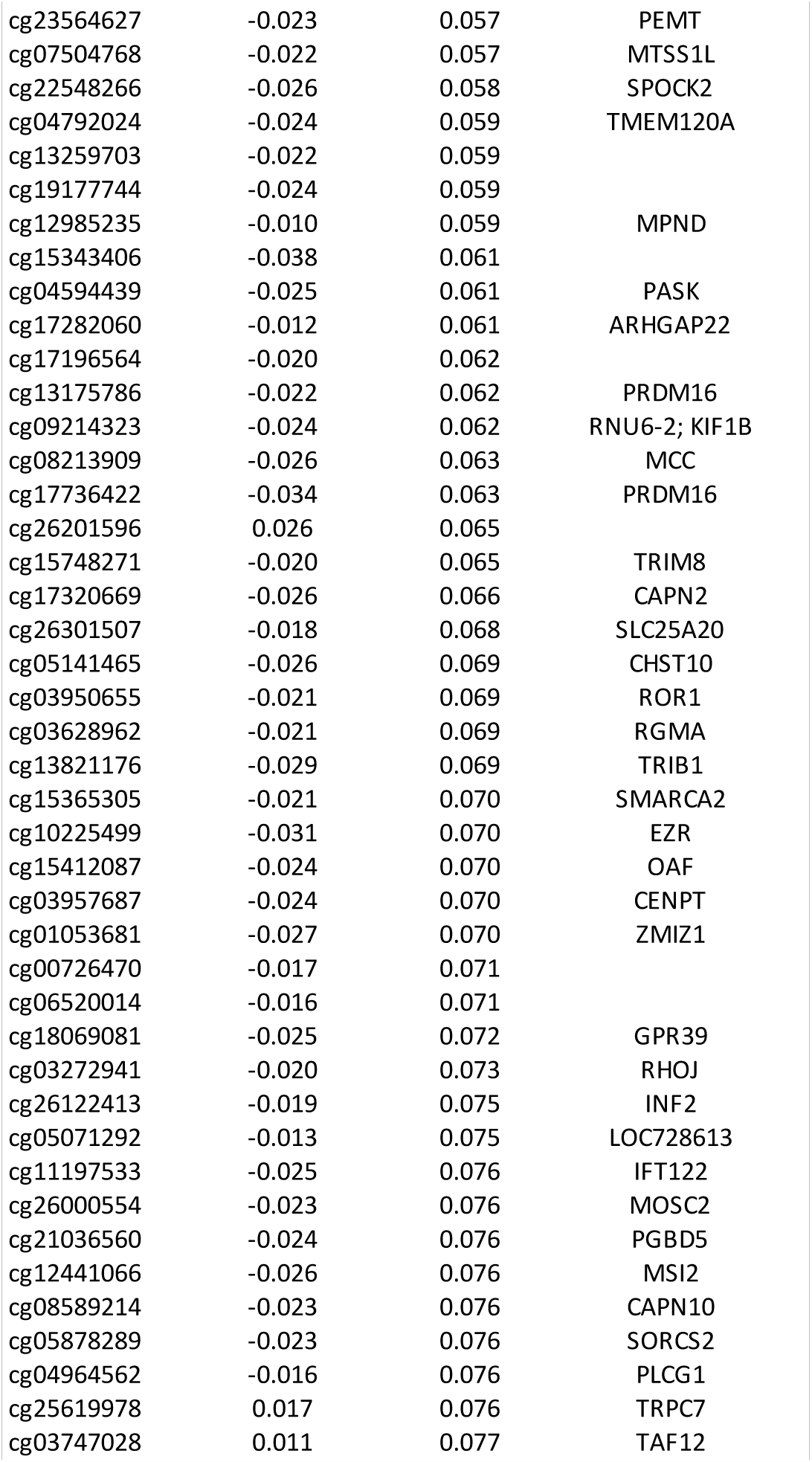
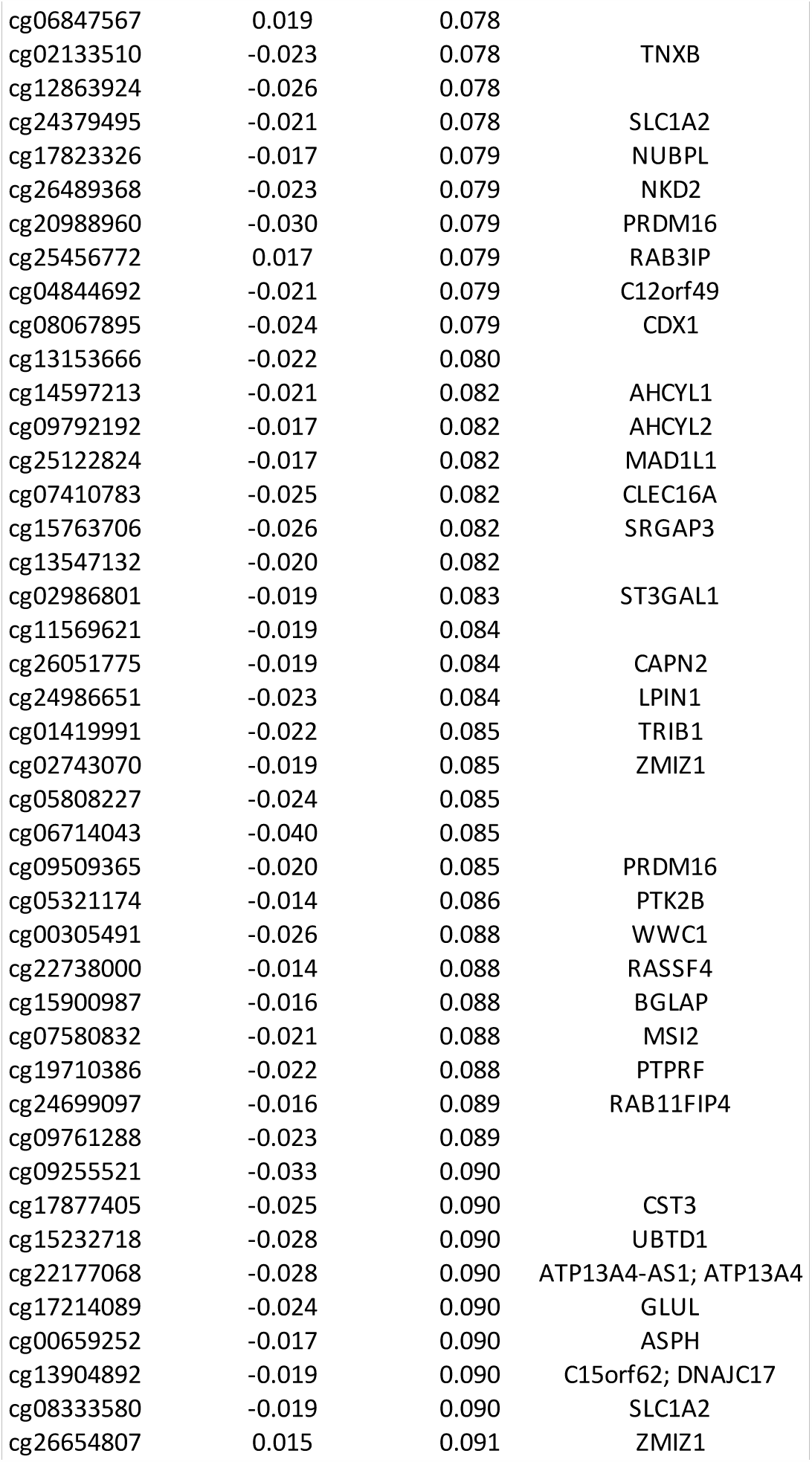
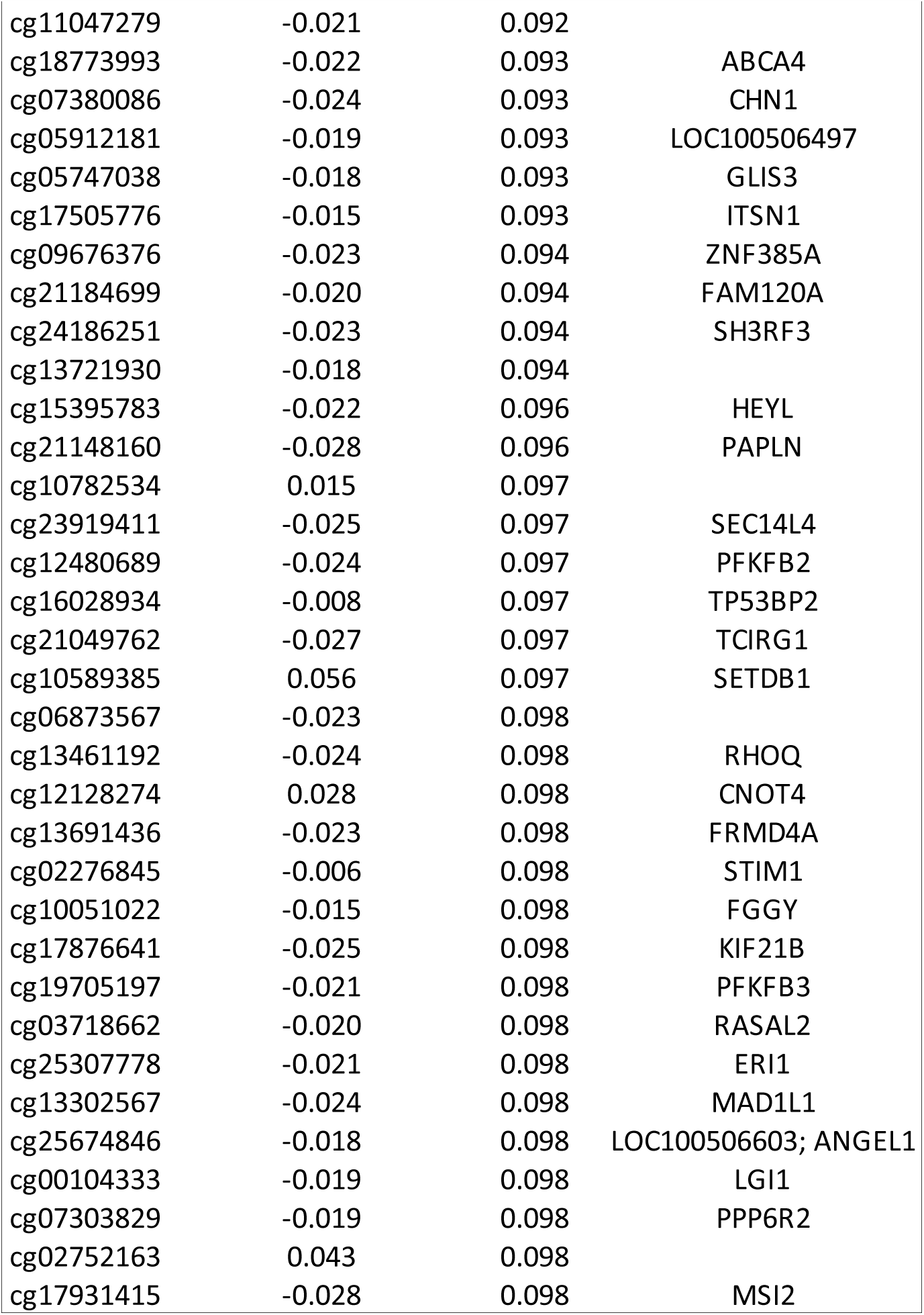
Differentially methylated sites in SZ. The 242 sites at which DNAm differed between SZ and NPC subjects with the FDR cutoff of q=0.1 (adjusted for age, race and PMI) are listed in the table. Abbreviations: DNAm, DNA methylation; NPC, non-psychiatric comparison; PMI, postmortem interval; SZ, schizophrenia.

DNAm is known to differ markedly between neurons and glia (36), and detection of DNAm differences between groups in tissue with multiple cell types can be confounded by cell composition. In STG samples studied here, neuronal proportion did not differ between subjects with SZ and NPC subjects (SZ=0.46 ±0.05; NPC=0.46 ±0.04; p=0.50) (Supplemental Table 2A), and we have previously shown that pyramidal neuron number in layer 3 of this brain region did not differ between subjects with SZ and NPC subjects (37). After adjusting for neuron proportion, DNAm differed at 256 sites between SZ and NPC subjects with the FDR cutoff of q=0.1 (Supplemental Table 2B). Of these 256 sites, 210 were among the 242 detected prior to adjusting for neuron proportion thus suggesting that cell composition does not account for the majority of observed differences in DNAm.

Genomic regions in which DNAm at multiple contiguous sites differs between SZ and NPC subjects, or differentially methylated regions (DMRs), may be more biologically meaningful or have different functional consequences than those of a single DMS. There were 44 genomic regions in which DNAm at two or more contiguous, measured sites differed between subjects with SZ and NPC subjects (Table 3).

**Table 3.** Differentially methylated regions in SZ. The 44 genomic regions in which DNAm differed at two or more contiguous, measured sites between SZ and NPC subjects are listed. Abbreviations: DNAm, DNA methylation; DMR, differentially methylated region; SZ, schizophrenia; NPC, non-psychiatric comparison; bp, base pairs.

| Chromosome | Start | End | Length in bp | Number of Significant Probes | Mean β-value Coefficient | Overlapping Promoters | Overlapping Genes |
| --- | --- | --- | --- | --- | --- | --- | --- |
| 6 | 28828946 | 28829503 | 557 | 15 | 0.012562318 | LINC01623, RPL13P | LINC01623, RPL13P |
| 1 | 16062361 | 16063471 | 1110 | 11 | -0.010632242 | SLC25A34, RP11-288I21.1 | SLC25A34, RP11-288I21.1 |
| 1 | 3191219 | 3192542 | 1323 | 10 | -0.021685336 |  | PRDM16 |
| 10 | 6263235 | 6264776 | 1541 | 9 | -0.016688209 | PFKFB3 | PFKFB3 |
| 19 | 48958216 | 48959043 | 827 | 9 | -0.014250403 | KCNJ14 | GRWD1 |
| 15 | 93617080 | 93617833 | 753 | 9 | -0.016227133 | RGMA | RGMA |
| 1 | 2999586 | 3001128 | 1542 | 8 | -0.020050418 |  | PRDM16 |
| 4 | 1800154 | 1801294 | 1140 | 8 | -0.012881307 | FGFR3 | FGFR3 |
| 3 | 187453721 | 187454786 | 1065 | 8 | -0.015068722 | BCL6 | BCL6 |
| 3 | 13082751 | 13083684 | 933 | 7 | -0.018337881 |  | IQSEC1 |
| 1 | 153600597 | 153600972 | 375 | 7 | -0.016748303 | S100A1, S100A13 | S100A1, S100A13 |
| 3 | 12949457 | 12950513 | 1056 | 6 | -0.015753296 |  | IQSEC1 |
| 4 | 1803298 | 1805090 | 1792 | 6 | -0.015286045 | FGFR3 | FGFR3 |
| 8 | 141588118 | 141588943 | 825 | 6 | -0.017229636 |  | AGO2 |
| 2 | 8711042 | 8711256 | 214 | 6 | -0.018511245 |  | LINC01814 |
| 4 | 1807819 | 1808646 | 827 | 6 | -0.01256232 | FGFR3 | FGFR3 |
| 4 | 866299 | 866811 | 512 | 6 | -0.017785773 | GAK | GAK |
| 14 | 21491808 | 21492316 | 508 | 6 | -0.010876489 | NDRG2 | NDRG2 |
| 10 | 99329961 | 99330447 | 486 | 5 | -0.014591035 | ANKRD2 | UBTD1 |
| 2 | 241535685 | 241536284 | 599 | 5 | -0.021733676 | CAPN10 | CAPN10 |
| 1 | 3154081 | 3154700 | 619 | 5 | -0.023621906 | PRDM16 | PRDM16 |
| 5 | 1033518 | 1034076 | 558 | 4 | -0.023946075 | NKD2 | NKD2 |
| 19 | 45411802 | 45412647 | 845 | 4 | -0.023104719 |  | APOE |
| 7 | 2262244 | 2262479 | 235 | 4 | -0.020981488 | MAD1L1 | MAD1L1 |
| 6 | 5087031 | 5087749 | 718 | 4 | -0.021469791 | PPP1R3G | PPP1R3G, LYRM4 |
| 9 | 138966848 | 138967347 | 499 | 4 | -0.021449114 |  | NACC2 |
| 10 | 7212815 | 7213304 | 489 | 4 | -0.021980366 |  | SFMBT2 |
| 12 | 117157450 | 117158154 | 704 | 4 | -0.015498341 | C12orf49 | C12orf49 |
| 10 | 3162054 | 3162191 | 137 | 4 | -0.017789142 |  | PFKP |
| 13 | 79170146 | 79170430 | 284 | 4 | 0.014049989 |  | OBI1-AS1 |
| 9 | 96199463 | 96199581 | 118 | 3 | -0.02791665 | RP11-165J3.6-001 |  |
| 17 | 55703709 | 55704098 | 389 | 3 | -0.029515629 |  | MSI2 |
| 5 | 167858238 | 167858326 | 88 | 3 | -0.017722017 | WWC1-008, WWC1-007 | WWC1 |
| 22 | 47016623 | 47017522 | 899 | 3 | -0.020100319 | GRAMD4 | GRAMD4 |
| 17 | 79686849 | 79687075 | 226 | 3 | -0.026164519 |  | SLC25A10 |
| 1 | 223945127 | 223945297 | 170 | 3 | -0.022033434 | CAPN2 | CAPN2 |
| 1 | 3128606 | 3129115 | 509 | 3 | -0.024216925 |  | PRDM16 |
| 15 | 93611859 | 93611950 | 91 | 2 | -0.024325293 |  | RGMA |
| 7 | 4829256 | 4829350 | 94 | 2 | -0.028189732 | AP5Z1, MIR4656 | AP5Z1 |
| 15 | 70364327 | 70364359 | 32 | 2 | -0.020032632 |  | TLE3 |
| 15 | 65594642 | 65594648 | 6 | 2 | -0.024798094 | PARP16 | RP11-349G13.2 |
| 14 | 65689711 | 65689836 | 125 | 2 | -0.021050816 | LINC02324 |  |
| 17 | 77954963 | 77955167 | 204 | 2 | -0.0173864 |  | TBC1D16 |
| 3 | 58558333 | 58558343 | 10 | 2 | -0.018019101 | RP11-475O23.2 | FAM107A |

Of note, three DMSs and one DMR were identified within mitotic arrest deficient 1-like 1 (*MAD1L1*), a gene strongly associated with SZ through genome-wide association studies (3). The *MAD1L1*-associated differential methylation we identified spanned a portion of intron 2 and exon 3, and the DNAm levels were lower in SZ subjects relative to NPC subjects.

### SZ-associated differential DNA methylation at some individual sites was specific to neurons or glia

Cell type deconvolution identified nine differentially methylated sites in neurons (Figure 2A and 2C) and two differentially methylated sites in glia (Figure 2B and 2C) with an FDR cutoff of q=0.1. All 11 sites for which DNAm differed between SZ and NPC subjects in cell type-specific manner were also identified as being differentially methylated in the bulk tissue analysis (Table 2).

**Figure 2.**
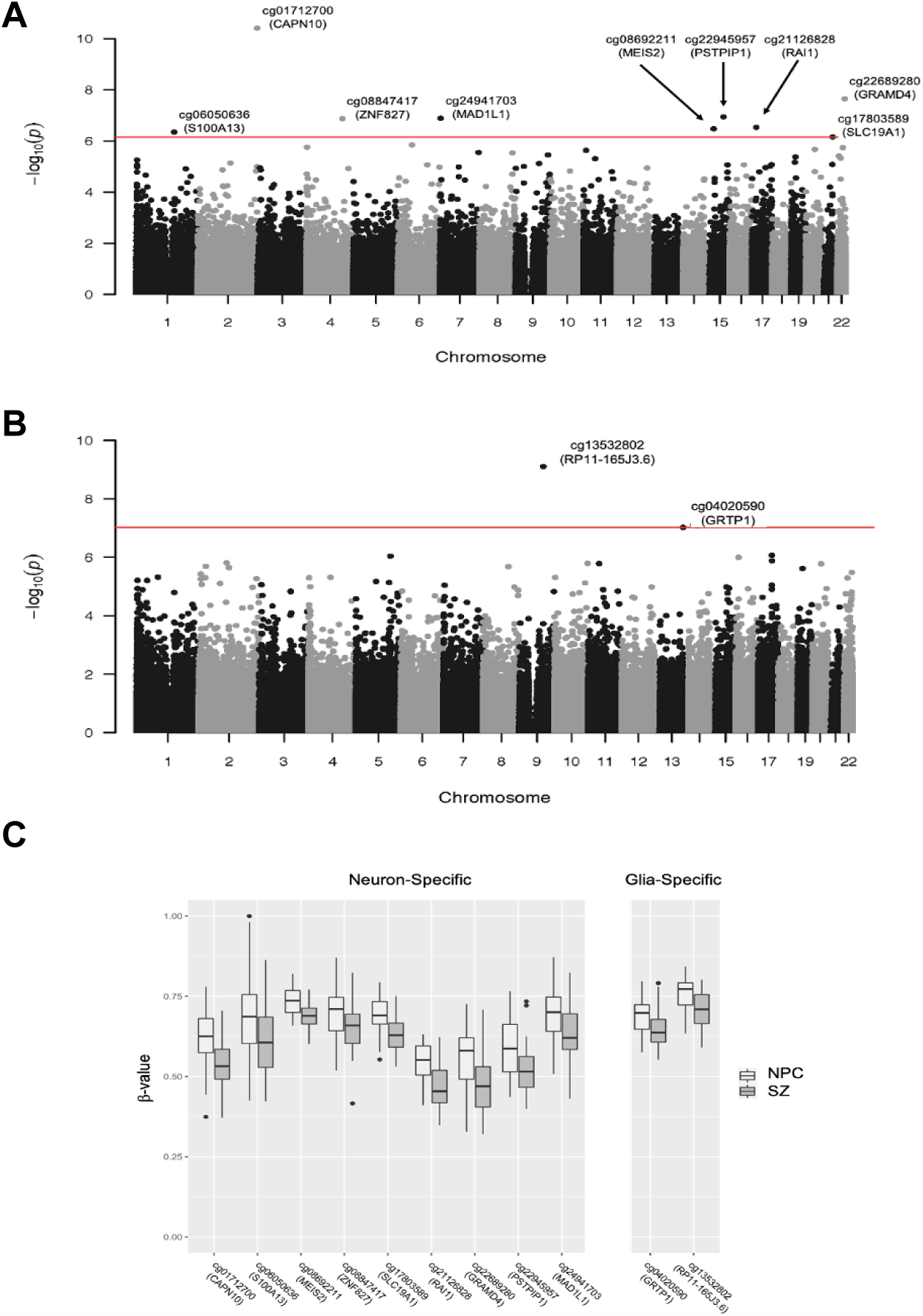
Neuron- or glia-specific differential methylation in SZ. **(A)** Manhattan plot showing neuron-specific DNAm differences between SZ and NPC subjects at nine sites. **(B)** Manhattan plot showing glia-specific DNAm differences between SZ and NPC subjects at two sites. **(C)** Box plots of DNAm (*β*-value) at sites of cell type-specific differences in DNAm between SZ and NPC subjects. DNAm, DNA methylation; SZ, schizophrenia; NPC, non-psychiatric comparison.

## DISCUSSION

In the STG of SZ subjects, we indentified DNAm differences at 242 individual sites and 44 genomic regions with multiple sites. Notably, we found SZ-associated differential methylation in *MAD1L1*, a gene that harbors a single nucleotide polymorphism (SNP) that strongly associates with SZ (third out of 145 loci that reached genome-wide significance) in the largest meta-analysis of genome-wide association studies to date from the Schizophrenia Working Group of the Psychiatric Genomics Consortium (3). The *MAD1L1*-associated differential methylation we identified was characterized by lower DNAm in SZ subjects relative to NPC subjects, a difference we determined to be driven by neuron-specific alterations in DNAm. This finding is consistent with studies in the prefrontal cortex that also identified genome-wide significant DMRs in MAD1L1 (10).

Despite strong evidence implicating *MAD1L1* in SZ, the underlying mechanisms through which MAD1L1-associated SNPs and differential methylation might contribute to SZ neurobiology are unclear. MAD1L1 is expressed in many human tissues (39,40), and is known to have a role in regulating the spindle assembly checkpoint during mitosis (41). Genetic mutations that disrupt MAD1L1 expression are associated with aneuploidy and multiple cancers (39,40). During development MAD1L1 is most strongly expressed in differentiating cells and is critical for the transition from proliferation to terminal differentiation in a broad range of cell types (42,43). Given MAD1L1 is expressed in both neurons and glia of most brain regions (44–46), the differentiation of neurons and glia may be disrupted if MAD1L1 expression is affected by SZ-associated differential methylation during neurodevelopment. Such a disruption would be predicted to alter the delicate balance of the various neuronal and glia subtypes and thus brain circuitry, perhaps giving rise to the dysfunctional brain circuits that are associated with the clinical features of SZ (47).

Alternatively, MAD1L1 may act post-neurodevelopment as its expression in terminally differentiated cells, including post-mitotic neurons and glia, suggests a function in addition to those related to development. Studies have found that its expression in terminally differentiated cells may be necessary for maintaining the differentiated state (48–50). Indeed, even modest decreases in MAD1L1 expression leads to dedifferentiation in some cell types (48). Some evidence points to a role for dedifferentiation of post-mitotic neurons in the cognitive decline and behavioral changes associated with normal brain aging in humans (51–53), and a similar mechanism could conceivably contribute to SZ neurobiology. That said, these proposed mechanisms are conjecture and critical next steps should focus on understanding MAD1L1 in the brain, generally, and translating MAD1L1-associated SNPs and differential methylation into molecular mechanisms for SZ, specifically.

This study is the first to identify SZ-associated differential methylation in the STG. Others have previously reported DNAm differences between subjects with SZ and NPC subjects in the prefrontal cortex (10,38,54–56), striatum (54), hippocampus (54,57), and cerebellum (54) thus suggesting that altered DNAm in multiple brain regions contributes to SZ neurobiology. Our findings add to the growing body of literature that implicates altered epigenetic pathways, including DNAm as well as histone modifications (58–60), in SZ neurobiology. Though most often studied separately, there is extensive crosstalk between DNAm and histone modification pathways (61–63). This crosstalk drives the establishment of composite epigenetic signatures that depend on epigenetic regulatory enzymes (e.g., DNA methyltransferases, histone methyltransferases, etc) with protein domains that specifically recognize methylated DNA and/or modified histones and thus allow for linking of DNAm and histone modifications at appropriate sites in the genome. SETD1A, a gene in which loss-of-function mutations confer a large increase in risk for SZ (64), is an example of an enzyme linking DNAm and histone modification. SETD1A methylates histones after it is localized to unmethylated DNA via an interaction with CXXC-finger protein-1 (65). This body of literature suggests that a more complete understanding of how these epigenetic pathways and their interactions are altered in SZ is likely to be fruitful in identifying molecular mechanisms contributing to SZ. Epigenetic editing technologies that use highly specific DNA-targeting tools (e.g., CRISPR) to methylate DNA or modify histones in a locus-specific manner will be valuable in dissecting these molecular mechanisms in cell culture and animal models (66,67).

Our findings in this study, like those of all postmortem brain studies, are only correlative and cannot establish causal relationships. The causes of the SZ-associated differences in DNAm that we identified in the STG are likely a combination of genetic and environmental factors. Methylation quantitative trait loci are common genetic variants that influence DNAm levels at sites in a particular genomic region, and they are common in the brain (68). However, none of the differentially methylated loci identified in this study have been shown to be associated with genetic variants that overlap with SZ risk loci (10,54), or are known to be altered by environmental risk factors for SZ like perinatal famine (11).

Though differential methylation may be associated with SZ risk factors, it may be the result of exposure to antipsychotics or others confounds. Studies of peripheral tissues indicate that antipsychotics do alter DNAm (69). However, DNAm alterations are already present in subjects with only brief (<16 weeks) antipsychotic treatment (70), thus suggesting that much SZ-associated differential methylation is intrinisic to the illness. Some studies have even found that SZ-associated DNAm alterations in peripheral tissues are normalized by treatment with antipsychotics (71), raising the possibility that the therapeutic effects of antipsychotics are mediated, in part, by DNAm changes. Such findings also make it likely that antipsychotics mask some SZ-associated differential methylation from being detected in studies.

An additional potential confound particularly relevant in studies of DNAm in subjects with SZ is cigarette smoking. Cigarette smoking is much more common among individuals with SZ than the general population and is known to induce robust DNAm changes in peripheral tissues (72). The effect of cigarette smoking on DNAm in the brain has not been studied. However, none of the differentially methylated sites or regions identified in this study have been found to be among the sites most strongly affected by cigarette smoking in peripheral tissues.

This study lays the groundwork for more detailed investigations of SZ-associated differential methylation in the STG. Future studies should focus identifying the mechanisms by which altered DNAm, especially within MAD1L1, contributes to SZ neurobiology. To this end, studies that use of epigenetic editing technology to recapitulate SZ-associated differential methylation in cell cultures and animal models will be useful. Also of value will be studies that use a multi-omics approach for understanding the dynamic interaction among epigenome, transcriptome, and genetic variation in the STG.

## Data Availability

Data is available for download from Gene Expression Omnibus (GEO; GSE144910)

https://www.ncbi.nlm.nih.gov/geo/query/acc.cgi?acc=GSE144910

## REFERENCES

1. Hilker R, Helenius D, Fagerlund B, Skytthe A, Christensen K, Werge TM, et al. Heritability of Schizophrenia and Schizophrenia Spectrum Based on the Nationwide Danish Twin Register. Biol Psychiatry. 2018;

2. Ripke S, Neale BM, Corvin A, Walters JTR, Farh KH, Holmans PA, et al. Biological insights from 108 schizophrenia-associated genetic loci. Nature. 2014;511(7510):421–7.

3. Pardiñas AF, Holmans P, Pocklington AJ, Escott-Price V, Ripke S, Carrera N, et al. Common schizophrenia alleles are enriched in mutation-intolerant genes and in regions under strong background selection. Nat Genet. 2018;

4. Genovese G, Fromer M, Stahl EA, Ruderfer DM, Chambert K, Landén M, et al. Increased burden of ultra-rare protein-altering variants among 4,877 individuals with schizophrenia. Nat Neurosci. 2016;

5. Marshall CR, Howrigan DP, Merico D, Thiruvahindrapuram B, Wu W, Greer DS, et al. Contribution of copy number variants to schizophrenia from a genome-wide study of 41,321 subjects. Nat Genet. 2017;

6. Van Os J, Rutten BPF, Poulton R. Gene-environment interactions in schizophrenia: Review of epidemiological findings and future directions. Schizophrenia Bulletin. 2008.

7. Tobi EW, Goeman JJ, Monajemi R, Gu H, Putter H, Zhang Y, et al. DNA methylation signatures link prenatal famine exposure to growth and metabolism. Nat Commun. 2014;

8. Heijmans BT, Tobi EW, Stein AD, Putter H, Blauw GJ, Susser ES, et al. Persistent epigenetic differences associated with prenatal exposure to famine in humans. Proc Natl Acad Sci U S A. 2008;

9. Hannon E, Spiers H, Viana J, Pidsley R, Burrage J, Murphy TM, et al. Methylation QTLs in the developing brain and their enrichment in schizophrenia risk loci. Nat Neurosci [Internet]. 2016;19(1):48–54. Available from: http://www.ncbi.nlm.nih.gov/pubmed/26619357

10. Jaffe AE, Gao Y, Deep-Soboslay A, Tao R, Hyde TM, Weinberger DR, et al. Mapping DNA methylation across development, genotype and schizophrenia in the human frontal cortex. Nat Neurosci [Internet]. 2016;19(1):40–7. Available from: http://www.ncbi.nlm.nih.gov/pubmed/26619358

11. Boks MP, Houtepen LC, Xu Z, He Y, Ursini G, Maihofer AX, et al. Genetic vulnerability to DUSP22 promoter hypermethylation is involved in the relation between in utero famine exposure and schizophrenia. npj Schizophr. 2018;

12. Jones PA. Functions of DNA methylation: Islands, start sites, gene bodies and beyond. Nature Reviews Genetics. 2012.

13. Wagner JR, Busche S, Ge B, Kwan T, Pastinen T, Blanchette M. The relationship between DNA methylation, genetic and expression inter-individual variation in untransformed human fibroblasts. Genome Biol. 2014;

14. Maunakea AK, Nagarajan RP, Bilenky M, Ballinger TJ, Dsouza C, Fouse SD, et al. Conserved role of intragenic DNA methylation in regulating alternative promoters. Nature. 2010;

15. Javitt DC, Sweet RA. Auditory dysfunction in schizophrenia: integrating clinical and basic features. Nat Rev Neurosci [Internet]. 2015;16(9):535–50. Available from: http://www.ncbi.nlm.nih.gov/pubmed/26289573

16. Sweet RA, Henteleff RA, Zhang W, Sampson AR, Lewis DA. Reduced dendritic spine density in auditory cortex of subjects with schizophrenia. Neuropsychopharmacology. 2009;34(2):374–89.

17. Glantz LA, Lewis DA. Decreased dendritic spine density on prefrontal cortical pyramidal neurons in schizophrenia. Arch Gen Psychiatry [Internet]. 2000;57(1):65–73. Available from: http://eutils.ncbi.nlm.nih.gov/entrez/eutils/elink.fcgi?dbfrom=pubmed&id=10632234&retmode=ref&cmd=prlinks%5Cnpapers3://publication/uuid/44227C59-6608-40A9-92C8-D27EF1BA4713

18. DSM-IV-TR. Diagnostic and statistical manual of mental disorders: DSM-IV-TR. DSM-V. 2000.

19. McKinney B, Ding Y, Lewis DA, Sweet RA. DNA methylation as a putative mechanism for reduced dendritic spine density in the superior temporal gyrus of subjects with schizophrenia. Transl Psychiatry [Internet]. 2017 Feb 14 [cited 2017 Jun 4];7(2):e1032. Available from: http://www.ncbi.nlm.nih.gov/pubmed/28195572

20. Nestler EJ, Peña CJ, Kundakovic M, Mitchell A, Akbarian S. Epigenetic Basis of Mental Illness. Neuroscientist. 2016;22(5):447–63.

21. Moore LD, L. T, Fan G. DNA methylation and its basic function. Neuropsychopharmacology. 2013 Jan;38(1):23–38.

22. Pidsley R, Zotenko E, Peters TJ, Lawrence MG, Risbridger GP, Molloy P, et al. Critical evaluation of the Illumina MethylationEPIC BeadChip microarray for whole-genome DNA methylation profiling. Genome Biol. 2016 Dec;17(1):208.

23. Moran S, Arribas C, Esteller M. Validation of a DNA methylation microarray for 850,000 CpG sites of the human genome enriched in enhancer sequences. Epigenomics [Internet]. 2016 Mar;8(3):389–99. Available from: http://www.ncbi.nlm.nih.gov/pubmed/26673039

24. Du P, Kibbe WA, Lin SM. lumi: a pipeline for processing Illumina microarray. Bioinformatics [Internet]. 2008;24(13):1547–8. Available from: http://www.ncbi.nlm.nih.gov/pubmed/18467348

25. Aryee MJ, Jaffe AE, Corrada-Bravo H, Ladd-Acosta C, Feinberg AP, Hansen KD, et al. Minfi: a flexible and comprehensive Bioconductor package for the analysis of Infinium DNA methylation microarrays. Bioinformatics. 2014 May;30(10):1363–9.

26. Cox TF, Cox MAA. Multidimensional scaling. Chapman & Hall/CRC; 2001. 308 p.

27. Xing J, Watkins WS, Shlien A, Walker E, Huff CD, Witherspoon DJ, et al. Toward a more uniform sampling of human genetic diversity: a survey of worldwide populations by high-density genotyping. Genomics [Internet]. 2010 Oct;96(4):199–210. Available from: http://www.ncbi.nlm.nih.gov/pubmed/20643205

28. Horvath S. DNA methylation age of human tissues and cell types. Genome Biol. 2013;

29. McKinney BC, Lin C-W, Rahman T, Oh H, Lewis DA, Tseng G, et al. DNA methylation in the human frontal cortex reveals a putative mechanism for age-by-disease interactions. Transl Psychiatry. 2019;9(1):39.

30. Akbarian S, Beeri MS, Haroutunian V. Epigenetic determinants of healthy and diseased brain aging and cognition. JAMA Neurol. 2013 Jun;70(6):711–8.

31. Lewis DA. The human brain revisited: opportunities and challenges in postmortem studies of psychiatric disorders. Neuropsychopharmacology [Internet]. 2002;26(2):143–54. Available from: http://www.ncbi.nlm.nih.gov/pubmed/11790510

32. McCullumsmith RE, Hammond JH, Shan D, Meador-Woodruff JH. Postmortem brain: an underutilized substrate for studying severe mental illness. Neuropsychopharmacology [Internet]. 2014 Jan;39(1):65–87. Available from: http://www.ncbi.nlm.nih.gov/pubmed/25767083

33. Peters TJ, Buckley MJ, Statham AL, Pidsley R, Samaras K, V Lord R, et al. De novo identification of differentially methylated regions in the human genome. Epigenetics and Chromatin. 2015;

34. Guintivano J, Aryee MJ, Kaminsky ZA. A cell epigenotype specific model for the correction of brain cellular heterogeneity bias and its application to age, brain region and major depression. Epigenetics [Internet]. 2013;8(3):290–302. Available from: http://www.ncbi.nlm.nih.gov/pubmed/23426267

35. Rahmani E, Schweiger R, Rhead B, Criswell LA, Barcellos LF, Eskin E, et al. Cell-type-specific resolution epigenetics without the need for cell sorting or single-cell biology. Nat Commun. 2019;

36. Kozlenkov A, Roussos P, Timashpolsky A, Barbu M, Rudchenko S, Bibikova M, et al. Differences in DNA methylation between human neuronal and glial cells are concentrated in enhancers and non-CpG sites. Nucleic Acids Res [Internet]. 2014 Jan;42(1):109–27. Available from: http://www.ncbi.nlm.nih.gov/pubmed/24057217

37. Dorph-Petersen KA, Delevich KM, Marcsisin MJ, Zhang W, Sampson AR, Gundersen HJG, et al. Pyramidal neuron number in layer 3 of primary auditory cortex of subjects with schizophrenia. Brain Res [Internet]. 2009;1285:42–57. Available from: http://www.ncbi.nlm.nih.gov/pubmed/19524554

38. Wockner LF, Noble EP, Lawford BR, Young RM, Morris CP, Whitehall VL, et al. Genome-wide DNA methylation analysis of human brain tissue from schizophrenia patients. Transl Psychiatry [Internet]. 2014;4:e339. Available from: http://www.ncbi.nlm.nih.gov/pubmed/24399042

39. Sun Q, Zhang X, Liu T, Liu X, Geng J, He X, et al. Increased expression of mitotic arrest deficient-like 1 (MAD1L1) is associated with poor prognosis and insensitive to taxol treatment in breast cancer. Breast Cancer Res Treat. 2013;

40. Tsukasaki K, Miller CW, Greenspun E, Eshaghian S, Kawabata H, Fujimoto T, et al. Mutations in the mitotic check point gene, MAD1L1, in human cancers. Oncogene. 2001;

41. Amon A. The spindle checkpoint. Curr Opin Genet Dev. 1999;

42. Hurlin PJ, Foley KP, Ayer DE, Eisenman RN, Hanahan D, Arbeit JM. Regulation of Myc and Mad during epidermal differentiation and HPV-associated tumorigenesis. Oncogene. 1995;

43. Ohta Y, Hamada Y, Saitoh N, Katsuoka K. Effect of the transcriptional repressor Mad1 on proliferation of human melanoma cells. Exp Dermatol. 2002;

44. Cichon S, Mühleisen TW, Degenhardt FA, Mattheisen M, Miró X, Strohmaier J, et al. Genome-wide association study identifies genetic variation in neurocan as a susceptibility factor for bipolar disorder. Am J Hum Genet. 2011;

45. Hawrylycz MJ, Lein ES, Guillozet-Bongaarts AL, Shen EH, Ng L, Miller JA, et al. An anatomically comprehensive atlas of the adult human brain transcriptome. Nature. 2012;

46. GTEx Project. GTEx portal. GTEx Analysis Release V6p (dbGaP Accession phs000424.v6.p1). 2017.

47. Lewis DA, Sweet RA. Schizophrenia from a neural circuitry perspective: advancing toward rational pharmacological therapies. J Clin Invest [Internet]. 2009;119(4):706–16. Available from: http://www.ncbi.nlm.nih.gov/pubmed/19339762

48. Han S, Park K, Kim HY, Lee MS, Kim HJ, Kim YD, et al. Clinical implication of altered expression of Mad1 protein in human breast carcinoma. Cancer. 2000;

49. Pan L, Zhang X, Suo X, Wang F, Niu Z, Dong Z. Expression and Mutation Analysis of the Myc Antagonist Gene Mad1 in Acute Leukemia. Blood. 2006;

50. Foley KP, Eisenman RN. Two MAD tails: What the recent knockouts of Mad1 and Mxi1 tell us about the MYC/MAX/MAD network. Biochimica et Biophysica Acta - Reviews on Cancer. 1999.

51. Oh G, Ebrahimi S, Wang SC, Cortese R, Kaminsky ZA, Gottesman II, et al. Epigenetic assimilation in the aging human brain. Genome Biol [Internet]. 2016;17:76. Available from: http://www.ncbi.nlm.nih.gov/pubmed/27122015

52. Koen JD, Rugg MD. Neural Dedifferentiation in the Aging Brain. Trends in Cognitive Sciences. 2019.

53. Park DC, Polk TA, Park R, Minear M, Savage A, Smith MR. Aging reduces neural specialization in ventral visual cortex. Proc Natl Acad Sci U S A. 2004;

54. Viana J, Hannon E, Dempster E, Pidsley R, Macdonald R, Knox O, et al. Schizophrenia-associated methylomic variation: molecular signatures of disease and polygenic risk burden across multiple brain regions. Hum Mol Genet [Internet]. 2017;26(1):210–25. Available from: http://www.ncbi.nlm.nih.gov/pubmed/28011714

55. Numata S, Ye T, Herman M, Lipska BK. DNA methylation changes in the postmortem dorsolateral prefrontal cortex of patients with schizophrenia. Front Genet [Internet]. 2014;5:280. Available from: http://www.ncbi.nlm.nih.gov/pubmed/25206360

56. Pai S, Li P, Killinger B, Marshall L, Jia P, Liao J, et al. Differential methylation of enhancer at IGF2 is associated with abnormal dopamine synthesis in major psychosis. Nat Commun. 2019;

57. Ruzicka WB, Subburaju S, Benes FM. Circuit- and Diagnosis-Specific DNA Methylation Changes at gamma-Aminobutyric Acid-Related Genes in Postmortem Human Hippocampus in Schizophrenia and Bipolar Disorder. JAMA Psychiatry [Internet]. 2015;72(6):541–51. Available from: http://www.ncbi.nlm.nih.gov/pubmed/25738424

58. O’dushlaine C, Rossin L, Lee PH, Duncan L, Parikshak NN, Newhouse S, et al. Psychiatric genome-wide association study analyses implicate neuronal, immune and histone pathways. Nat Neurosci. 2015;

59. Chase KA, Rosen C, Rubin LH, Feiner B, Bodapati AS, Gin H, et al. Evidence of a sex-dependent restrictive epigenome in schizophrenia. J Psychiatr Res. 2015;

60. Fullard JF, Halene TB, Giambartolomei C, Haroutunian V, Akbarian S, Roussos P. Understanding the genetic liability to schizophrenia through the neuroepigenome. Schizophrenia Research. 2016.

61. Hashimoto H, Vertino PM, Cheng X. Molecular coupling of DNA methylation and histone methylation. Epigenomics. 2010.

62. Du J, Johnson LM, Jacobsen SE, Patel DJ. DNA methylation pathways and their crosstalk with histone methylation. Nat Rev Mol Cell Biol. 2015;

63. Ooi SKT, Qiu C, Bernstein E, Li K, Jia D, Yang Z, et al. DNMT3L connects unmethylated lysine 4 of histone H3 to de novo methylation of DNA. Nature. 2007;

64. Singh T, Kurki MI, Curtis D, Purcell SM, Crooks L, McRae J, et al. Rare loss-of-function variants in SETD1A are associated with schizophrenia and developmental disorders. Nat Neurosci. 2016;

65. Tate CM, Lee JH, Skalnik DG. CXXC finger protein 1 restricts the Setd1A histone H3K4 methyltransferase complex to euchromatin. FEBS J. 2010;

66. Holtzman L, Gersbach CA. Editing the Epigenome: Reshaping the Genomic Landscape. Annu Rev Genomics Hum Genet. 2018;

67. Stricker SH, Köferle A, Beck S. From profiles to function in epigenomics. Nature Reviews Genetics. 2016.

68. Gibbs JR, van der Brug MP, Hernandez DG, Traynor BJ, Nalls MA, Lai SL, et al. Abundant quantitative trait loci exist for DNA methylation and gene expression in human brain. PLoS Genet [Internet]. 2010;6(5):e1000952. Available from: http://www.ncbi.nlm.nih.gov/pubmed/20485568

69. Castellani CA, Melka MG, Diehl EJ, Laufer BI, O’Reilly RL, Singh SM. DNA methylation in psychosis: insights into etiology and treatment. Epigenomics [Internet]. 2015;7(1):67–74. Available from: http://www.ncbi.nlm.nih.gov/pubmed/25687467

70. Nishioka M, Bundo M, Koike S, Takizawa R, Kakiuchi C, Araki T, et al. Comprehensive DNA methylation analysis of peripheral blood cells derived from patients with first-episode schizophrenia. J Hum Genet [Internet]. 2013;58(2):91–7. Available from: http://www.ncbi.nlm.nih.gov/pubmed/23235336

71. Abdolmaleky HM, Pajouhanfar S, Faghankhani M, Joghataei MT, Mostafavi A, Thiagalingam S. Antipsychotic drugs attenuate aberrant DNA methylation of DTNBP1 (dysbindin) promoter in saliva and post-mortem brain of patients with schizophrenia and Psychotic bipolar disorder. Am J Med Genet Part B Neuropsychiatr Genet. 2015;

72. Joehanes R, Just AC, Marioni RE, Pilling LC, Reynolds LM, Mandaviya PR, et al. Epigenetic Signatures of Cigarette Smoking. Circ Cardiovasc Genet. 2016;9(5):436–47.

